# Can the ChatGPT and other Large Language Models with internet-connected database solve the questions and concerns of patient with prostate cancer?

**DOI:** 10.1101/2023.03.06.23286827

**Authors:** Lingxuan Zhu, Weiming Mou, Rui Chen

**Affiliations:** Department of Urology, Ren Ji Hospital, Shanghai Jiao Tong University School of Medicine, Shanghai, 200127, China; The First Clinical Medical School, Southern Medical University, 1023 Shatai South Road, Guangzhou, 510515, Guangdong, China

## Abstract

Large language models (LLMs), such as ChatGPT, have shown impressive natural language processing capabilities in various fields, including medicine. However, the answers provided by these models may sometimes be incorrect, and they may not have access to the latest data. In this study, we aimed to evaluate the performance of five state-of-the-art LLMs in providing correct and comprehensive information on common questions raised by prostate cancer patients. We also examined whether LLMs with internet-connected databases could provide more up-to-date information than ChatGPT. We designed a set of 22 questions covering various aspects of prostate cancer and evaluated the accuracy, comprehensiveness, patient readability, and inclusion of humanistic care in the answers provided by each model. Our findings suggest that although the performance of different LLMs varied, these LLMs could provide accurate basic knowledge and have the ability to analyze specific situations to a certain extent. We also found that the overall performance of the LLM model with internet-connected dataset was not superior to ChatGPT, and the paid version of ChatGPT did not show superiority over the free version. Our study highlights the potential of LLMs in bridging the gap between patients and healthcare providers. Current LLMs have the potential to be applied for patient education and consultation, providing patient-friendly information. Shared decision-making with the doctors and patients could be achieved easier. We believed that with the rapid development of AI technology, LLMs have unlimited potential.

## Introduction & Methods

Large language models (LLMs) represented by ChatGPT have demonstrated remarkable abilities in natural language processing. In the medical field, ChatGPT can pass the USMLE exam^1^ and provide advice on preventing cardiovascular diseases to patients^2^. However, it should be noted that the answers provided by ChatGPT may contain many errors, and OpenAI also stated that ChatGPT may give answers that appear correct but are incorrect^3^. In addition, ChatGPT is only trained based on data before September 2021 and cannot access the latest data on the Internet. In the months following ChatGPT’s launch, OpenAI released a paid version, ChatGPT Plus, to handle high traffic volumes. Other companies have also launched LLMs with internet-connected database to compete with ChatGPT. These models can answer the questions based on the latest information on the Internet and therefore the answer may have better timeliness. Taking prostate cancer as an example, we evaluated whether these LLMs could offer correct and useful information on common questions raised by patients with prostate cancer or patient who concerns with prostate cancer and provide appropriate humanistic care. We aimed to evaluate whether these LLMs can help bridge the gap between patients and healthcare providers.

Prostate cancer is the second-most common type of cancer in men globally, with relatively a long survival time compared with other cancer types^4^. The widespread use of prostate specific antigen (PSA) screening has generated many suspected prostate cancer patients, and if diagnosed, the treatment process from detection to castrate-resistant prostate cancer (CRPC) or metastasis is also very lengthy, which has created a large demand for consultations. We designed a set of 22 questions (Table 1) based on patient education guidelines from institutions such as GDC and UpToDate, as well as our own clinical experience. These questions cover various aspects of prostate cancer, including screening, prevention, treatment options, and postoperative complications. The difficulty level ranges from basic knowledge to specific situational analysis and cutting-edge knowledge of prostate cancer diagnosis and treatment. A total of five state-of-the-art LLMs were included, including ChatGPT (Feb 13 Free Version & Plus version Default model), YouChat, NeevaAI, Perplexity (concise & detailed model), and Chatsonic. Except for NeevaAI and Perplexity, the other models generated different responses each time, so we generated three responses for each question in these models to examine the stability of the models. The quality of the answers was primarily evaluated based on their accuracy, using a 3-point scale: 1 for correct, 2 for mixed with correct and incorrect/outdated data, and 3 for completely incorrect. The answers were further evaluated for comprehensiveness, patient readability (on a 5-point Likert scale), and whether they included humanistic care for the patient (yes or no). The internal consistency of each model’s answers was evaluated after review.

**Table 1.**
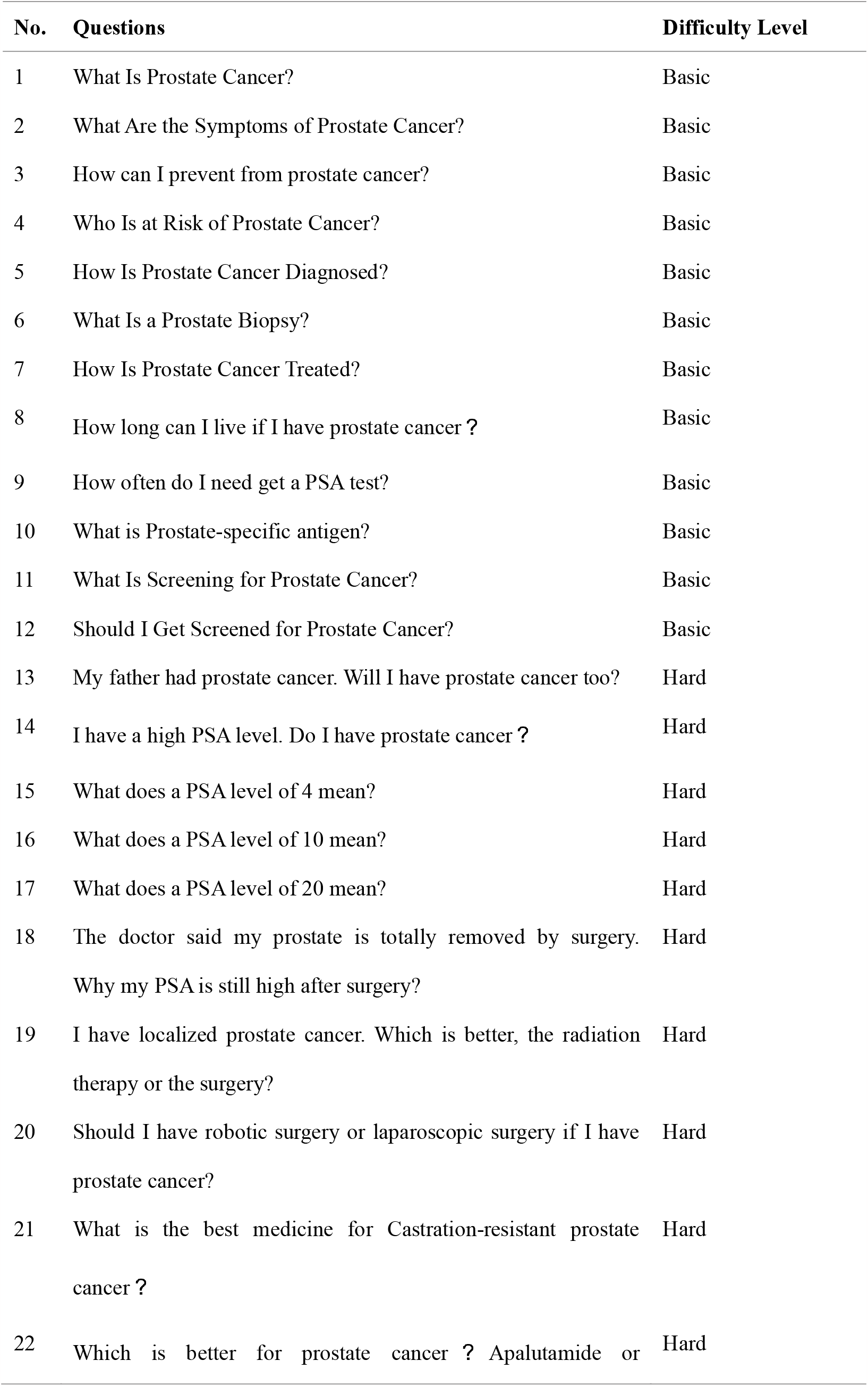
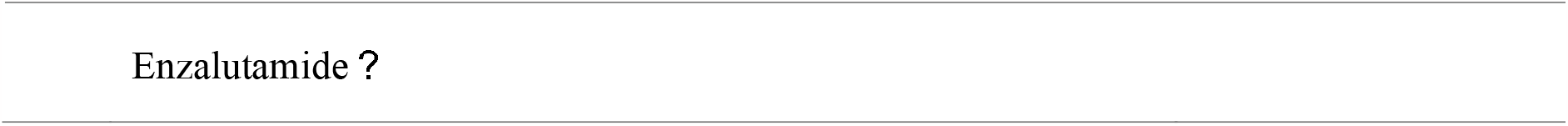
Questions and corresponding difficulty levels used to test the performance of LLMs.

| No. | Questions | Difficulty Level |
| --- | --- | --- |
| 1 | What Is Prostate Cancer? | Basic |
| 2 | What Are the Symptoms of Prostate Cancer? | Basic |
| 3 | How can I prevent from prostate cancer? | Basic |
| 4 | Who Is at Risk of Prostate Cancer? | Basic |
| 5 | How Is Prostate Cancer Diagnosed? | Basic |
| 6 | What Is a Prostate Biopsy? | Basic |
| 7 | How Is Prostate Cancer Treated? | Basic |
| 8 | How long can I live if I have prostate cancer ? | Basic |
| 9 | How often do I need get a PSA test? | Basic |
| 10 | What is Prostate-specific antigen? | Basic |
| 11 | What Is Screening for Prostate Cancer? | Basic |
| 12 | Should I Get Screened for Prostate Cancer? | Basic |
| 13 | My father had prostate cancer. Will I have prostate cancer too? | Hard |
| 14 | I have a high PSA level. Do I have prostate cancer ? | Hard |
| 15 | What does a PSA level of 4 mean? | Hard |
| 16 | What does a PSA level of 10 mean? | Hard |
| 17 | What does a PSA level of 20 mean? | Hard |
| 18 | The doctor said my prostate is totally removed by surgery. Why my PSA is still high after surgery? | Hard |
| 19 | I have localized prostate cancer. Which is better, the radiation therapy or the surgery? | Hard |
| 20 | Should I have robotic surgery or laparoscopic surgery if I have prostate cancer? | Hard |
| 21 | What is the best medicine for Castration-resistant prostate cancer ? | Hard |
| 22 | Which is better for prostate cancer ? Apalutamide or | Hard |

We aimed to evaluate: 1) Whether LLMs can be used as a consultant for prostate cancer patients. 2) Whether LLMs can answer the question associated with the latest developments in prostate cancer manage. 3) Whether the paid version of ChatGPT is superior to the free version. 4) Whether the LLMs with internet-connected database could provide more up-to-date information compared with ChatGPT.

## Result

LLMs could provide correct answers to the questions of concern in prostate cancer patient. The accuracy of most LLMs’ responses was above 90%, except for NeevaAI and Chatsonic (83.33% and 75%, respectively). For basic information questions with definite answers (such as “How Is Prostate Cancer Diagnosed?”, “What Is Prostate Cancer?”), most LLMs could achieve a high accuracy. Nevertheless, the accuracy decreased in questions associated with specific scenario, or in questions that involved summary and analysis (e.g., Why the PSA is still high after surgery?). Among these LLMs, ChatGPT had the highest accuracy rate, and the free version of ChatGPT was slightly better than the paid version. The main reason for poor answer accuracy was mixing correct and incorrect or outdated information in the responses.

We further evaluated the comprehensiveness of the answer provided by LLMs. ChatGPT has the highest comprehensiveness score among all LLMs, while the comprehensiveness of other models is less satisfactory. The comprehensiveness score of the Perplexity Concise was lower than that of Perplexity Detailed. LLMs perform well in answering most questions. For example, they can effectively point out the significance of different PSA levels for patients and correctly remind patients that PSA is not the final diagnostic test and further examination is necessary. In terms of comparing treatment options, most LLMs can accurately and comprehensively state the advantages and disadvantages of two methods, providing a reference for patients’ choices. In addition, it is commendable that most responses point out the need for patients to consult their doctors for more advice.

We examined the readability of the responses of each LLM to evaluate whether they could be understood by patients when potentially applied in clinical settings. The readability of responses from most LLMs, except NeevaAI, was satisfactory, and we believe that patients can understand the information conveyed in LLM responses in most cases.

We then analyzed the reasons for the poor performance of LLMs in some responses. In regard to the question about PSA elevation after prostatectomy, most LLMs did not mention that recurrence or metastasis could cause an increase in PSA levels. NeevaAI incorrectly suggested that postoperative trauma could lead to PSA elevation. In some responses, LLMs failed to understand the background information of the question and provided inaccurate answers, such as indicating that postoperative PSA elevation was due to prostatitis or infection even when the patient had already undergone prostatectomy, and in another response, mechanically suggested that “PSA testing is not the final diagnostic test for prostate cancer,” but monitoring PSA after prostatectomy is clearly not for the purpose of diagnosing prostate cancer. Mixed outdated or incorrect information included claiming that compared to robotic-assisted surgery (currently the most common option for radical prostatectomy^5^), laparoscopic surgery is the more common choice, and that laparoscopic surgery is a traditional surgical procedure that requires a large incision for access to the prostate. The main reason for inadequate comprehensiveness was the lack of specific details or omission of key points. For example, Perplexity missed screening as an important measure in preventing prostate cancer. In the question of how often one should undergo PSA testing, some responses only stated that it should be analyzed on a case-by-case basis, lacking specific details on testing frequency for different age groups^6^. In terms of readability, NeevaAI and the detail mode of Perplexity sometimes provided irrelevant information, which affected readability. It must be noted that some AI models based on search engines such as NeevaAI tend to simply provide the content of literature without summarizing and explaining, leading to poor readability.

For the question related to a specific condition to choose one medication from two (Which is better for prostate cancer? Apalutamide or Enzalutamide?) Most of the LLMs failed to give satisfied responses. For these questions, the LLMs should first of all ask the enquirer additional questions such as the stage and status, yet all LLMs failed. The major failure of the LLMs were they could not give accurate responses about the approved indications of both medications. Both of them were approved for non-metastatic castration-resistant prostate cancer and metastatic castration-sensitive prostate cancer, however, only Enzalutamide was approved for metastatic castration-resistant prostate cancer. Other mistakes of the LLMs in answering this question included failure in giving information of one medication, giving outdated information (such as meeting abstract in 2018), and providing irrelevant information about other medications (Darolutamide).

Finally, we evaluated the performance of the LLM in terms of humanistic care and its stability over multiple responses. All LLMs demonstrated humanistic care only when answering questions about expected lifespan, and most LLM were able to provide patients with correct information that the survival time of prostate cancer is relatively long, which can help alleviate their anxiety. However, they did not demonstrate any humanistic care when answering other questions. In terms of stability, the LLM was mostly consistent, but inconsistent results were observed in a few cases.

## Discussion

Using prostate cancer as an example, our findings further illustrate the promising application of LLMs in patient education and medical consultation. Although not yet perfect, the vast majority of LLMs can provide correct answers to basic questions that prostate cancer patients are concerned about and have the ability to analyze specific situations to a certain extent. Additionally, the responses provided by LLMs are generally correct, comprehensive, helpful, and easy to understand. Among these models, ChatGPT performs the best in all aspects, with the most detailed descriptions and the most accessible language. It also frequently emphasizes that it is an AI and suggests that patients seek further help from medical professionals. Our research also found that the paid version of ChatGPT does not offer superior answer quality compared to the free version, therefore, patients do not need to purchase the paid version.

In terms of response quality, the accuracy assessment stratified by question difficulty illustrates that LLMs can answer the basics about the disease well. The main reasons affecting accuracy errors due to the inclusion of outdated information in the responses and failure to understand the contextual information of complex questions. Also, our assessment of the comprehensiveness of the responses pointed out that LLMs sometimes missed some key points and may lack specific details in the responses. While we anticipated that the internet-connected LLM model would surpass ChatGPT, we discovered that it did not outperform ChatGPT overall. Current internet-connected LLM models tend to rely on simple merging and integration of search content on top of traditional search engines, sometimes simply providing literature conclusions rather than fully reflecting the complete answer to a question, resulting in poor readability. This suggests that model training may be relatively more critical. Our research also highlights the serious lack of humanistic care in current LLMs, which cannot proactively comfort anxious patients in their responses. Therefore, current LLMs are not yet capable of completely replace doctors, as they may contain errors or omit key points in responses and still have significant shortcomings in analyzing questions in specific contexts. Moreover, they still cannot comfort patients like humans.

In summary, although the performance of different LLMs varied, it was shown that these LLMs could provide accurate basic knowledge. Current LLMs have the potential to be applied for patient education and consultation, providing patient-friendly information. Shared decision-making with the doctors and patients could be achieved easier. Additionally, LLMs have the ability to analyze specific situations to a certain extent, making them potentially useful for simple online consultations in the future. We believed that with the rapid development of AI technology, LLMs have unlimited potential.

We acknowledged the limitations of this study. As these models are still in the testing phase, it is reasonable to believe that their accuracy may improve with continuous iteration and optimization. Moreover, we only tested common issues regarding a specific disease. Testing more complex issues with multiple diseases may be more comprehensive. Nonetheless, we believed our research indicated the feasibility of LLMs for future doctor-patient communication. The results of this study should be treated carefully although it indeed shed light upon the possible future of AI-based healthcare.

## Data Availability

The data that support the findings of this study are available on request from the corresponding author upon reasonable request.

## Statements & Declarations Funding

This study is supported by the Rising-Star Program of Science and Technology Commission of Shanghai Municipality (21QA1411500), Natural Science Foundation of Science and Technology Commission of Shanghai (22ZR1478000), and the National Natural Science Foundation of China (82272905)

## Competing Interests

The authors declare that the research was conducted in the absence of any commercial or financial relationships that could be construed as a potential conflict of interest.

## CRediT Author Contributions

**LZ:** Conceptualization, Methodology, Investigation, Formal analysis, Writing - Original Draft, Visualization**; WM:** Visualization, Writing - Review & Editing**; RC:** Supervision, Funding acquisition, Writing - Review & Editing, Conceptualization, Methodology,

## Figure legdens

**Figure 1.**
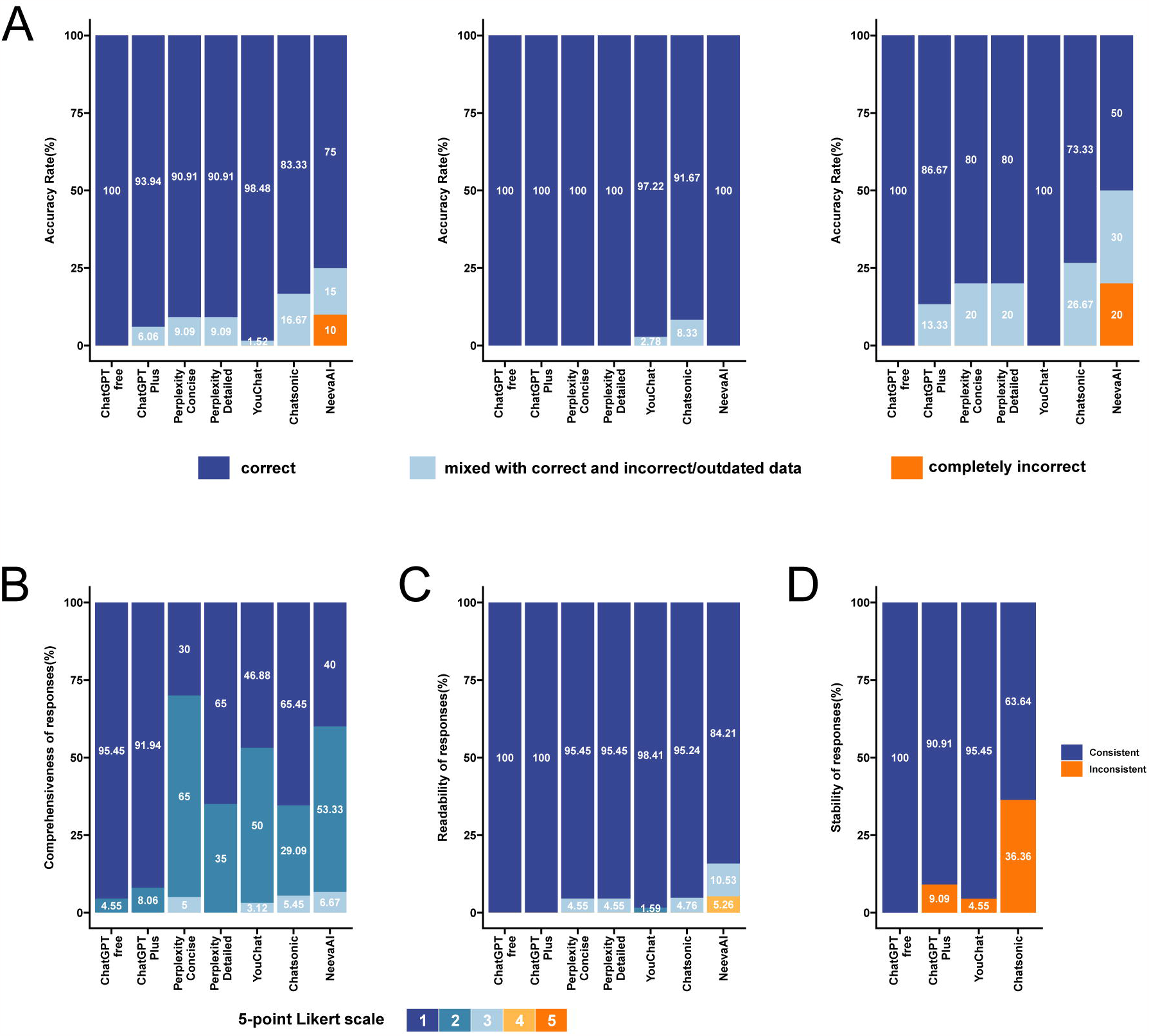
The performance of several large language models (LLMs) in answering different questions. **A**. Accuracy of responses. From left to right, the performance in all questions, the performance in basic questions, and the performance in difficult questions. **B**. The comprehensiveness of correctly answered responses. A 5-point Likert scale is used, with 1 representing “very comprehensive” and 5 representing “very Inadequate”. **C**. Readability of answers. A 5-point Likert scale is used, with 1 representing “very easy to understand” and 5 representing “very difficult to understand”. **D**. Stability of responses. Judged based on whether the model’s accuracy is consistent across different responses to the same question.

## Notes

### Competing Interest Statement

The authors have declared no competing interest.

